# The cry as a vital sign: A smartphone based study of neonatal cry acoustics in hypoxic ischemic encephalopathy

**DOI:** 10.64898/2026.10.12.26365334

**Authors:** Debankur Chakraborty

## Abstract

**Background:** Many Indian special newborn care units (SNCUs) cannot offer amplitude-integrated EEG (aEEG) or early MRI, so bedside judgement decides which newborns receive closer neurological attention. Earlier work found that asphyxiated newborns cry with a higher pitch and shorter phonation than healthy newborns [10], and a meta-analysis reported higher fundamental frequency, dysphonation and atypical melody in infants with neurological dysfunction, with wide differences between studies [11].

**Aim:** To determine whether basic acoustic features of a smartphone-recorded pain cry (following a heel prick) can differentiate term newborns with HIE from control newborns. Also, to track changes in cry acoustics over the first 72 hours of life and evaluate their association with the clinical severity and short-term outcomes of encephalopathy.

**Design:** A prospective, single-centre diagnostic accuracy and cohort study in a tertiary SNCU. Three groups of term newborns are recorded: newborns with HIE, healthy comparators, and sick comparators who do not have HIE. Every newborn is recorded after the same stimulus, the first clinically indicated heel-prick. Newborns with HIE and sick comparators are recorded again at 24 hours, 72 hours and before discharge. A cry that is late or absent counts as a result and not as a missing value. Three clinicians blinded to group also classify the cries by listening, so the phone-based measures can be compared with the human ear.

**Analysis:** The primary outcome is the area under the ROC curve of a logistic model with five pre-specified acoustic predictors. The target is 180 analysable newborns (70 with HIE and 110 comparators). Reporting follows STARD and TRIPOD+AI.

**Relevance:** In a South Asian trial, cooling did not reduce death or disability at 18 months and increased death alone [7]. A cry tool in this setting is therefore proposed to help prioritise EEG, seizure surveillance, closer observation and referral. It is not proposed as a trigger for cooling.

## 1. Introduction

Intrapartum related neonatal encephalopathy remains a major cause of neonatal death and impairment worldwide [1]. Severity is usually graded with the Sarnat stages [2] or a modified version of them, and with the Thompson score [3]. In one cohort, the Thompson score at 3 to 5 hours of age was a sensitive predictor of an abnormal aEEG at 6 hours [4]. Cooling trials began treatment within 6 hours of birth [5,6], so the first hours mainly decide what can be offered.

The HELIX trial randomised 408 newborns with moderate or severe encephalopathy in India, Sri Lanka and Bangladesh. Cooling did not reduce death or disability at 18 months but increased death alone, and the authors advised against offering it in these settings [7]. This changes what an early cry tool should aim to do. In this protocol its purpose is to flag newborns who need priority for EEG or a seizure surveillance, closer observation and early referral.

A cry combines breathing with laryngeal control, and the lower brainstem is thought to coordinate both [8,9]. Variation in fundamental frequency (F0, the basic pitch) has been linked to brainstem control of laryngeal tension through the vagal complex and the phrenic and thoracic nerves. Instability in these pathways is thought to produce hyperphonation, a sudden shift to an F0 above 1000 Hz [9].

Michelsson and colleagues analysed 115 pain cries from 45 term newborns with perinatal asphyxia and compared them with cries from 75 healthy newborns. The asphyxiated group had shorter phonation, a higher maximum and minimum pitch, and more of biphonation. The cries were more abnormal in children later found to be neurologically damaged [10]. A systematic review and meta-analysis of infants up to 18 months old with or at risk of neurological dysfunction found higher F0, dysphonation and atypical melodies [11]. Spectral features of cry audio have also been examined in relation to HIE [12], and an Indian doctoral thesis reported classification of HIE cries [13].

On the machine-learning side, one study applied neural transfer learning to a pre-recorded cry database to detect perinatal asphyxia [14], and a smartphone application for birth attendants has been proposed [15]. A review of cry analysis and classification is available [16]. Smartphone recordings made in uncontrolled settings with background noise have supported classification of pain cries in infants aged 0 to 24 months [17]; but that study did not include newborns with encephalopathy. Reviews describe acoustic cry analysis as a way to add an objective measure to listener perception [18], which varies with the listener’s sleep and mental state [17].

Seven features of the design sets this protocol apart from a simple comparison of HIE cries with heel-prick cries:

1. One stimulus for every newborn. All newborns are recorded after the same type of stimulus, the first clinically indicated heel-prick.
2. A comparator that matches the clinical question. Healthy newborns are an easy comparison. Sick newborns who do not have HIE, such as those with suspected sepsis or symptomatic hypoglycaemia, are the babies a nurse might confuse with HIE.
3. The cry over time. A single 15-second recording is a snapshot. Encephalopathy evolves over the first days [2], so the cry is recorded again at 24 hours, 72 hours and before discharge.
4. Silence counts as data. Infants in Sarnat stage 3 are stuporous and flaccid with suppressed brainstem function [2], and some may cry late, weakly or not at all. Dropping them would remove the group where a cry tool matters most, so cry status and latency are outcomes.
5. Pain intensity is measured. Cry pitch and structure change with pain intensity in term newborns [19], and both cry and facial expression carry pain information [20]. A bedside Neonatal Infant Pain Scale score [21] is recorded so that differences in pain response are not read as differences in brain injury.
6. The ear is tested against the phone. Three blinded clinicians classify the same clips, so the value added by the acoustic measures is judged against usual practice.
7. Recording under ward conditions. Recordings are made as in routine care. Ward noise is logged, a hand-held spacer fixes the distance, and two phone models are checked against each other.

## 2. Aim, objectives and hypotheses

### 2.1 Aim

To test whether simple acoustic measures from a smartphone recording of the cry after a heel-prick separate term newborns with HIE from other newborns, and whether the course of the cry over the first 72 hours follows the severity and outcome of the encephalopathy.

### 2.2 Primary objective

To determine whether basic acoustic features of a smartphone-recorded pain cry (following a heel prick) can differentiate term newborns with HIE from control newborns.

### 2.3 Secondary objectives

1. To compare each cry measure between the HIE group, healthy comparators and sick comparators.
2. To relate the measures to the modified Sarnat stage and the Thompson score at the time of recording.
3. To describe the change in the cry from the first recording to 72 hours and to test whether it relates to death or an abnormal neurological examination at discharge.
4. To estimate how often the cry is late or absent in each group and at each Sarnat stage.
5. To compare classification by the phone-based measures with classification by blinded clinicians who listen to the same clips.
6. To check agreement of the measures between two phone models and the effect of ward noise.

### 2.4 Hypothesis

Compared with newborns without HIE, newborns with HIE will have a higher first-cry F0, more hyperphonation, more dysphonation and shorter cry sounds [10,11]. The size of the deviation will increase with Sarnat stage. Two further hypotheses are exploratory: a cry that stays far from the comparator range at 72 hours will be associated with death or an abnormal examination at discharge, and a late or absent cry will be more frequent at stage 3 than at stage 2. No numerical F0 cut-offs are fixed in advance, apart from the 1000 Hz definition of hyperphonation [9]. Cut-offs will come from the data.

## 3. Methods

### 3.1 Design and setting

This will be a prospective, single-centre, observational study in the SNCU of a tertiary hospital. Enrolment can run for 9 months or until the target is reached. The study adds no procedure to routine care. Reporting will follow STARD [22] for diagnostic accuracy and TRIPOD+AI [23] for the prediction model. The protocol will be registered with the Clinical Trials Registry of India (CTRI) before enrolment.

### 3.2 Participants

All newborns are term (37 weeks or more) and would be enrolled after written parental consent. Table 1 shows the three groups. Group assignment follows the reference examination in section 3.4 and not the admission diagnosis, so a newborn admitted with suspected HIE who is later found not to have HIE moves to the sick comparator group.

**Table 1.** Study groups and recording schedule.

| Group | Who | Recordings |
| --- | --- | --- |
| HIE | History of perinatal asphyxia (Apgar below 5 at 5 minutes, or positive-pressure ventilation) and encephalopathy on examination by a clinician. | First recording within 6 hours of life where possible, then 24 hours, 72 hours and before discharge |
| Healthy comparators | No encephalopathy, routine heel-prick indicated; matched to the HIE group on gestational age (within 1 week) and age at recording (within 12 hours) | One recording |
| Sick comparators | Another condition that can make a newborn irritable or lethargic, for example suspected sepsis or meningitis, symptomatic hypoglycaemia, or seizures with a cause other than HIE; no HIE on examination | Same schedule as the HIE group |

Exclusion criteria are known vocal cord paralysis, cleft lip or palate, congenital diaphragmatic hernia, major airway or brain malformation, and intubation at the time of recording.

### 3.3 Recording procedure

- Equipment: two current smartphone models (one Android, one iPhone) with a free voice-recorder app set to uncompressed WAV at 44.1 kHz. Airplane mode is on and cloud backup is off.
- Position: the microphone is held 30 cm from the newborn’s mouth, using a 30 cm spacer taped to the phone. One trained staff member makes the recordings wherever possible.
- Stimulus: the first routine heel-prick for glucose monitoring. No extra pricks are made for the study. Recording starts 10 seconds before the prick and continues for 60 seconds after it.
- Timing: the first recording in the HIE group is made within 6 hours of life where possible and before any phenobarbitone or other sedative or anticonvulsant. If a drug was given earlier, its name, dose and time are logged.
- Later recordings: at 24 hours and 72 hours (each give or take 6 hours) and before discharge, a heel-prick is used if one is due. Otherwise, any cry during routine handling is recorded, and the stimulus type is logged.
- Context: age in hours, time of last feed, sucrose or other analgesia, cooling status, and ward noise level from a free sound-meter app over the 10 seconds before the prick. A bedside nurse scores the Neonatal Infant Pain Scale [21] within 1 minute of the prick.
- Device check: in the first 30 newborns, a second phone model records at the same time from the same distance, on the other side of the spacer.

If a newborn does not cry within 10 seconds, the recording continues and the outcome is logged as a late cry or, if no sound occurs within 60 seconds, as no cry. These newborns stay in the study.

### 3.4 Clinical assessment and outcomes

A paediatrician who has not heard the recording examines each HIE and sick-comparator newborn within 1 hour of each recording and assigns the modified Sarnat stage [2] and the Thompson score [3]. Healthy comparators are examined once. Cranial ultrasound, EEG are noted when done in routine care but are not required. Outcomes at discharge are death and the Hammersmith Neonatal Neurological Examination optimality score [24], assessed by an examiner who is blind to the acoustic data. The cut-off for an abnormal examination will be set before analysis. A standardised neurological examination at 3 months will be added if resources allow.

### 3.5 Acoustic measures

The first 30 seconds of cry after the prick are analysed. Measures are extracted with free software such as openSMILE [17], using one script that is fixed before data collection starts. Cry sounds are marked automatically and a second person checks the marking on the spectrogram. The analyst does not know the group. Table 2 lists the measures.

**Table 2.**
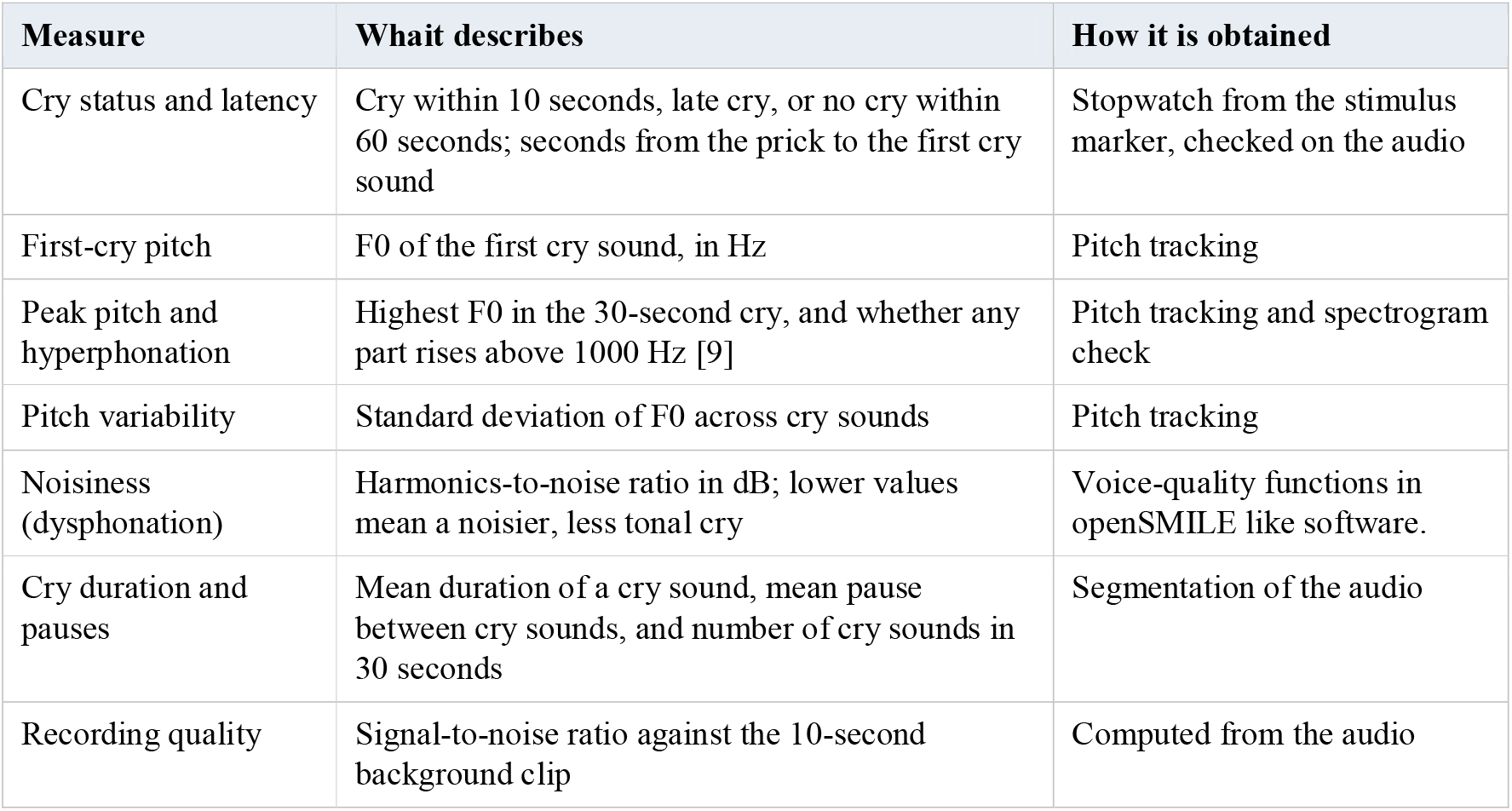
Acoustic measures.

Mel-spectrograms and Mel-frequency cepstral coefficients (MFCCs) are also computed. They are used only for exploratory clustering and do not enter the primary model.

### 3.6 Listening panel

Three listeners (a professor, resident and SNCU medical officer) classify the clips from the first recording. Clips are 10 seconds long, trimmed to remove speech, shuffled, and played on headphones. For each clip the listener chooses ‘normal’, ‘pain only’ or ‘neurologically abnormal’, and gives a confidence score from 0 to 10. Listeners do not know the group or the acoustic results.

## 4. Analysis plan and sample size

### 4.1 Primary analysis

The primary model is a logistic regression of HIE versus no HIE (healthy and sick comparators combined). The acoustic predictors are first-cry F0, F0 variability, harmonics-to-noise ratio, cry sound duration, and hyperphonation (yes or no). Age at recording and the pain scale score are added as adjustment variables. The model is limited to these seven parameters so that it stays within what 70 events can support, and the sample size will be checked against the criteria in reference [25].

Newborns who make no sound stay in the analysis. A no-cry indicator is added to the model and their missing acoustic values are replaced by the overall mean, a standard missing-indicator approach. A sensitivity analysis restricted to newborns who cried is also reported. Performance is reported as the area under the ROC curve with a bootstrap 95% confidence interval, corrected for optimism, together with a calibration plot, the sensitivity at 90% specificity and the specificity at 90% sensitivity. The same model is also tested on HIE versus sick comparators alone, which is the harder question.

### 4.2 Secondary and exploratory analyses

- Group comparison of each measure with ANOVA or the Kruskal-Wallis test, with pairwise follow-up tests.
- Trend across Sarnat stages with ordinal logistic regression or Spearman correlation.
- Change over time with a mixed model that has a random intercept for each newborn and a time-by-group term. The association of the change in F0 between the first recording and 72 hours with death or an abnormal discharge examination is tested with logistic regression.
- Listening panel: accuracy, sensitivity and specificity for each listener, kappa between listeners, and a paired bootstrap comparison of the panel’s AUC with the model’s AUC.
- Phone agreement: Bland-Altman plots and the intraclass correlation for F0 and harmonics-to-noise ratio. Ward noise: signal-to-noise ratio added as a covariate, with a test of its interaction with group.
- Exploratory: principal component analysis of the MFCCs with a two-dimensional scatter plot, and clustering without group labels to see whether clusters follow group or follow ward noise.

### 4.3 Sample size

The target is 70 analysable newborns with HIE and 110 comparators (55 healthy and 55 sick), 180 in total. With these numbers, a two-sided test at the 0.05 level has 90% power to detect a standardised difference of 0.5 in a single acoustic measure between HIE and all comparators, and 97% power for a difference of 0.6. If the model’s AUC is 0.85, the 95% confidence interval is about 0.79 to 0.91, using a standard approximation for the variance of the AUC. For the listening panel, 180 clips and a true kappa of 0.6 give a 95% confidence interval of about plus or minus 0.12, assuming an even split between classes. The effect sizes are assumptions chosen to represent a moderate difference and are not taken from a published study of this design.

To allow for 15% unusable recordings, the plan is to enrol 82 newborns with HIE and 64 in each comparator group, 210 in all. Recruitment depends on how many term newborns with HIE the unit admits. The investigator will check the admission register for the previous 12 months before the start. The plan assumes about 9 HIE admissions a month, and enrolment will be extended to 12 months if fewer are expected.

## 5. Ethics, privacy and data sharing

The study needs approval from the institutional ethics committee before enrolment. Written informed consent is sought from a parent or guardian before the first recording. If parents cannot be reached in time, the first recording is skipped and not made without consent. No extra heel-pricks are made, and recording never delays resuscitation or treatment. Clinicians do not see acoustic results during the study, and management does not change because of them.

Recordings can capture parents’ and staff voices. Speech is removed before clips are archived. Audio is stored on an encrypted drive under a study code, and the code key is kept separately. Phones used for recording have cloud backup switched off. After publication, the de-identified table of acoustic measures and clinical variables and the analysis script will be shared in a public repository. Trimmed audio clips will be available on request under a data-sharing agreement.

## 6. Expected results and limitations

We expect the HIE group to show higher and more variable pitch, more hyperphonation and noisier cries, in the direction found in earlier work [10,11]. We do not expect a clean split. Cry is variable between newborns, and overlap with sick comparators is likely. The study is designed to measure that overlap. If sick comparators sit close to the HIE group, the cry will not be useful as a triage signal even when HIE differs from healthy newborns.

The main limitations are these:

- The study is single-centre, and a model built on 70 events needs external validation before any clinical use.
- The reference standard is clinical. Without required MRI or EEG, some newborns will be misclassified.
- A heel-prick is a mild stimulus, and results may not hold for spontaneous cry. Spontaneous cries recorded at later time points are analysed separately for this reason.
- Drugs, sucrose, cooling and age at recording can change the cry. They are recorded but cannot be fully controlled.
- Few newborns at stage 3 are expected, so stage-specific estimates will be wide.
- The study cannot show that using the tool improves outcomes.

## 7. Dissemination

Results will be reported following STARD [22] and TRIPOD+AI [23] and submitted to a neonatology or paediatric neurology journal. The extraction script and the de-identified feature table will be published with the paper. Findings will be shared with SNCU staff and, in plain language, with parents of enrolled newborns.

## Data Availability

All data produced in the present work are contained in the manuscript

## References

1. Lee AC, Kozuki N, Blencowe H, Vos T, Bahalim A, Darmstadt GL, et al. Intrapartum-related neonatal encephalopathy incidence and impairment at regional and global levels for 2010 with trends from 1990. Pediatr Res. 2013;74 Suppl 1:50–72. doi:10.1038/pr.2013.206

2. Sarnat HB, Sarnat MS. Neonatal encephalopathy following fetal distress: a clinical and electroencephalographic study. Arch Neurol. 1976;33(10):696–705. doi:10.1001/archneur.1976.00500100030012

3. Thompson CM, Puterman AS, Linley LL, Hann FM, van der Elst CW, Molteno CD, et al. The value of a scoring system for hypoxic ischaemic encephalopathy in predicting neurodevelopmental outcome. Acta Paediatr. 1997;86(7):757–61. doi:10.1111/j.1651-2227.1997.tb08581.x

4. Horn A, Swingler G, Myer L, Linley L, Raban M, Joolay Y, et al. Early clinical signs in neonates with hypoxic ischemic encephalopathy predict an abnormal amplitude-integrated electroencephalogram at age 6 hours. BMC Pediatr. 2013;13:52. doi:10.1186/1471-2431-13-52

5. Jacobs SE, Berg M, Hunt R, Tarnow-Mordi WO, Inder TE, Davis PG. Cooling for newborns with hypoxic ischaemic encephalopathy. Cochrane Database Syst Rev. 2013;(1):CD003311. doi:10.1002/14651858.CD003311.pub3

6. Shankaran S, Laptook AR, Ehrenkranz RA, Tyson JE, McDonald SA, Donovan EF, et al. Whole-body hypothermia for neonates with hypoxic-ischemic encephalopathy. N Engl J Med. 2005;353(15):1574–84. doi:10.1056/NEJMcps050929

7. Thayyil S, Pant S, Montaldo P, Shukla D, Oliveira V, Ivain P, et al. Hypothermia for moderate or severe neonatal encephalopathy in low-income and middle-income countries (HELIX): a randomised controlled trial in India, Sri Lanka, and Bangladesh. Lancet Glob Health. 2021;9(9):e1273–85. doi:10.1016/S2214-109X(21)00264-3

8. Golub HL, Corwin MJ. A physioacoustic model of the infant cry. In: Lester BM, Boukydis CFZ, editors. Infant crying: theoretical and research perspectives. New York: Plenum; 1985. p. 59–82.

9. Zeskind PS, McMurray MS, Cox Lippard ET, Grewen KM, Garber KA, Johns JM. Translational analysis of effects of prenatal cocaine exposure on human infant cries and rat pup ultrasonic vocalizations. PLoS One. 2014;9(10):e110349. doi:10.1371/journal.pone.0110349

10. Michelsson K, Sirvio P, Wasz-Hockert O. Pain cry in full-term asphyxiated newborn infants correlated with late findings. Acta Paediatr Scand. 1977;66:611–6. doi:10.1111/j.1651-2227.1977.tb07956.x

11. Lawford HLS, Sazon H, Richard C, Robb MP, Bora S. Acoustic cry characteristics of infants as a marker of neurological dysfunction: a systematic review and meta-analysis. Pediatr Neurol. 2022;129:72–9. doi:10.1016/j.pediatrneurol.2021.10.017

12. Satar M, Cengizler C, Hamitoglu S, Ozdemir M. Investigation of relation between hypoxic-ischemic encephalopathy and spectral features of infant cry audio. J Voice. 2022. doi:10.1016/j.jvoice.2022.05.015

13. Chittora A. A signal processing based approach for infant cry analysis and classification [doctoral thesis]. Gandhinagar: Dhirubhai Ambani Institute of Information and Communication Technology; 2016.

14. Onu CC, Lebensold J, Hamilton WL, Precup D. Neural transfer learning for cry-based diagnosis of perinatal asphyxia. In: Proceedings of Interspeech 2019. 2019. p. 3053–7. doi:10.21437/Interspeech.2019-2340

15. Onu CC, Udeogu I, Ndiomu E, Kengni U, Precup D, Sant′anna GM, et al. Ubenwa: cry-based diagnosis of birth asphyxia. arXiv:1711.06405 [Preprint]. 2017.

16. Ji C, Mudiyanselage TB, Gao Y, Pan Y. A review of infant cry analysis and classification. EURASIP J Audio Speech Music Process. 2021;2021:8. doi:10.1186/s13636-021-00197-5

17. Parga JJ, Lewin S, Lewis J, Montoya-Williams D, Alwan A, Shaul B, et al. Defining and distinguishing infant behavioral states using acoustic cry analysis: is colic painful? Pediatr Res. 2020;87(3):576–80. doi:10.1038/s41390-019-0592-4

18. LaGasse LL, Neal AR, Lester BM. Assessment of infant cry: acoustic cry analysis and parental perception. Ment Retard Dev Disabil Res Rev. 2005;11:83–93. doi:10.1002/mrdd.20050

19. Bellieni CV, Sisto R, Cordelli DM, Buonocore G. Cry features reflect pain intensity in term newborns: an alarm threshold. Pediatr Res. 2004;55(1):142–6. doi:10.1203/01.PDR.0000099793.99608.CB

20. Grunau RV, Craig KD. Pain expression in neonates: facial action and cry. Pain. 1987;28:395–410. doi:10.1016/0304-3959(87)90073-X

21. Lawrence J, Alcock D, McGrath P, Kay J, MacMurray SB, Dulberg C. The development of a tool to assess neonatal pain. Neonatal Netw. 1993;12:59–66.

22. Bossuyt PM, Reitsma JB, Bruns DE, Gatsonis CA, Glasziou PP, Irwig L, et al. STARD 2015: an updated list of essential items for reporting diagnostic accuracy studies. BMJ. 2015;351:h5527. doi:10.1136/bmj.h5527

23. Collins GS, Moons KGM, Dhiman P, Riley RD, Beam AL, Van Calster B, et al. TRIPOD+AI statement: updated guidance for reporting clinical prediction models that use regression or machine learning methods. BMJ. 2024;385:e078378. doi:10.1136/bmj-2023-078378

24. Dubowitz L, Mercuri E, Dubowitz V. An optimality score for the neurologic examination of the term newborn. J Pediatr. 1998;133(3):406–16.

25. Riley RD, Ensor J, Snell KIE, Harrell FE Jr, Martin GP, Reitsma JB, et al. Calculating the sample size required for developing a clinical prediction model. BMJ. 2020;368:m441. doi:10.1136/bmj.m441

